# Risk Model for Intraoperative Complication during Cataract Surgery Based on Data from 900,000 Eyes – Previous Intravitreal Injection is a Risk Factor

**DOI:** 10.1101/2020.12.08.20245738

**Authors:** Poya Hård af Segerstad

**Author notes:** CORRESPONDING AUTHOR Poya Hård af Segerstad, Skåne University Hospital, Ophthalmology, Dept. of Clinical Sciences, Lund, Lund University, Klinikgatan 18, SE-221 85 Lund, SWEDEN, ADDRESS FOR REPRINTS: Corresponding author.

## Abstract

**Background/Aims:** The aim of this study was to develop a risk model for intraoperative complication during cataract surgery and to include previous intravitreal therapy in the model.

**Methods:** This retrospective register-based study covered patients reported to the Swedish National Cataract Register (SNCR) between Jan. 1, 2010 and Jun. 30, 2018. Odds ratios (OR) were used to quantify association strength of each variable with intraoperative complication. Data from the SNCR were cross referenced with the Swedish Macula Register (SMR) to include data on previous intravitreal therapy. Variables statistically significant in the univariate analyses (*P* <0.05) were included in a multivariate logistic regression model.

**Results:** The inclusion criteria were met by 907,499 eyes. The overall rate of intraoperative complication was 0.86%. After cross referencing, 3,451 eyes were identified in the SMR as having undergone intravitreal therapy prior to cataract surgery. Variables significantly associated with intraoperative complication (*P*<0.05) were best corrected visual acuity ≥1.0 LogMAR (adjusted OR): 1.75), age ≥90 years (OR: 1.25), male sex (OR: 1.09), pseudoexfoliation (OR: 1.33), glaucoma (OR: 1.11), diabetic retinopathy (OR: 1.35), previous intravitreal therapy (OR: 1.45), surgeon’s experience <600 surgeries (OR: 2.77), use of rhexis hooks (OR: 6.14), blue staining (OR: 1.87), and mechanical pupil dilation (OR: 1.52).

**Conclusion:** The risk model can be used in the preoperative setting to predict the probability of intraoperative complication, to facilitate planning of surgery, and improving patient communication. Patients who have undergone intravitreal therapy prior to cataract surgery have an increased risk of intraoperative complication during cataract surgery.

## INTRODUCTION

Cataract is one of the most common causes of visual impairment, with an estimated global prevalence of 17.2%^1^. The procedure of cataract surgery, phacoemulsification, is known to be safe with a low complication rate^2-4^, and it is therefore difficult to identify factors that may lead to intraoperative complication (IC). Despite the low rate of complications, additional surgical procedures may be required when they do occur, compromising both patient satisfaction and the final visual outcome^5^. Large dataset studies are therefore required to identify factors associated with IC^4, 6^, and previous intravitreal injection has recently been identified as a risk factor^7, 8^. Intravitreal injections of anti-vascular endothelial growth factor (anti-VEGF) are mainly performed as treatment for choroidal neovascularization in eyes with age-related macular degeneration^9^. With an increasingly aged population in Western countries, the number of cataract surgeries and intravitreal injections can be expected to increase^10-12^. The Swedish National Cataract Register (SNCR) includes data on patients who have undergone cataract surgery in Sweden since its inception in 1992, and has a high coverage rate^13^. In 2010, the Swedish personal ID number was included in the register, making it possible to cross reference it with data in other databases^14^. Similarly, the Swedish Macula Register (SMR) contains data on eyes subjected to treatment for choroidal neovascularization, collected since 2008^15^.

The primary aim of this study was to develop a risk model for IC during cataract surgery, based on data from 907,499 eyes reported to the SNCR. The model could be used in the preoperative clinical setting to facilitate planning of cataract surgery and improving patient communication. The secondary aim was to cross reference the dataset from SNCR with the Swedish Macula Register (SMR) to include data on previous intravitreal therapy (pIVT) in the model.

## MATERIALS AND METHODS

Data on cataract surgery performed between January 1, 2010 and June 30, 2018 were extracted from the SNCR^13^. The following data were considered relevant and were used in this study: Swedish personal ID number, surgeon’s ID, date of surgery, patient’s right or left eye, sex, preoperative best-corrected visual acuity (BCVA), axial length, comorbidities (pseudoexfoliation, glaucoma, macular degeneration, diabetic retinopathy, corneal endothelial degeneration, uveitis, previous refractive surgery, and previous vitrectomy), intraoperative difficulties (use of rhexis hooks, blue color staining and mechanical pupil dilation), indication for surgery (poor visual acuity, anisometropia, elevated intraocular pressure (IOP), other visual disturbance and “other”), and whether communication occurred between the anterior and posterior segments during surgery, referred to as “intraoperative complication” in this study. Data on intravitreal therapy with no date restrictions (Swedish personal ID number, date of injection and right or left eye) were extracted from the SMR^15^. Data on procedures other than phacoemulsification and patients younger than 20 years at time of surgery were excluded. The SNCR data center assisted in calculating the patient’s age at the time of cataract surgery and age at the time of intravitreal therapy from the datasets. As the collection of surgeon’s ID was introduced in the SNCR from 2007, the data center also assisted in calculating the accumulated number of surgeries performed per surgeon from 2007 and onward. They also anonymized the data by replacing the personal ID number and the ID of the cataract surgeon with randomized numbers. The data from each register were cross referenced and eyes that had received intravitreal therapy prior to cataract surgery were identified. Visual acuity reported as Snellen chart scores was converted to Logarithm of the Minimum Angle of Resolution (LogMAR) prior to analysis using the formula *LogMAR* = -*log*_10_ (*Snellen*)^16^. Infinite LogMAR values derived from Snellen scores of 0.00 were rounded to 2.0 (*n*=34).

### Statistical analysis

To identify cut-off levels at which each continuous variable (age, BCVA, surgeon’s experience and axial length) significantly affected the risk of IC, the variables were divided into intervals: 10 years for age, 0.1 for LogMAR, 200 surgeries and 1 mm in axial length. The odds ratio (OR) of IC was then calculated as the ratio between the odds of IC within each interval and the odds of IC outside each interval, before the variables were converted into discrete variables and divided at the level found to affect risk outcome (odds ratio ≥1.5). Odds ratios were used to quantify association strength in order to estimate the univariate association of each variable with IC. Nominal variables statistically significant in the univariate analyses (*P* <0.05) were included in a multivariate logistic regression model. Variables found to be nonsignificant in the multivariate analysis were removed from the model. Odds ratios were converted into probability to make them easier to use in the clinical setting. The study was approved by the Ethics Committee of Lund University, and was conducted according to the principles described in the Declaration of Helsinki. All statistical analyses were performed using R^17^.

## RESULTS

The inclusion criteria were met by 907,499 eyes in 572,536 patients in the SNCR. After cross referencing, 35,739 injections of 3,451 eyes in 3,168 patients were identified in the SMR as having undergone intravitreal therapy prior to cataract surgery. The mean age at the time of cataract surgery was 74.4 years, the proportions of male eyes and patients were 40.3% and 58.9%, respectively. The mean age at the time of intravitreal therapy was 81.0 years, the proportions of male eyes and patients were 37.3% and 37.9%, respectively. Patient characteristics and missing data are presented in Table 1. The amount of missing data was generally low, apart from the variables axial length, pseudoexfoliation, uveitis, previous refractive surgery, previous vitrectomy and indications for surgery, as these data were collected during limited periods. The continuous variables surgeon’s experience and BCVA were dichotomized at 600 surgeries and 1.0 LogMAR (corresponding to a Snellen score of 0.10), respectively, while age was trichotomized at 40 years and 90 years. Axial length was found to be nonsignificant for all values and was therefore excluded from further analyses. The level-to-level odds ratio comparisons are presented in Figure 1.

**Table 1.**
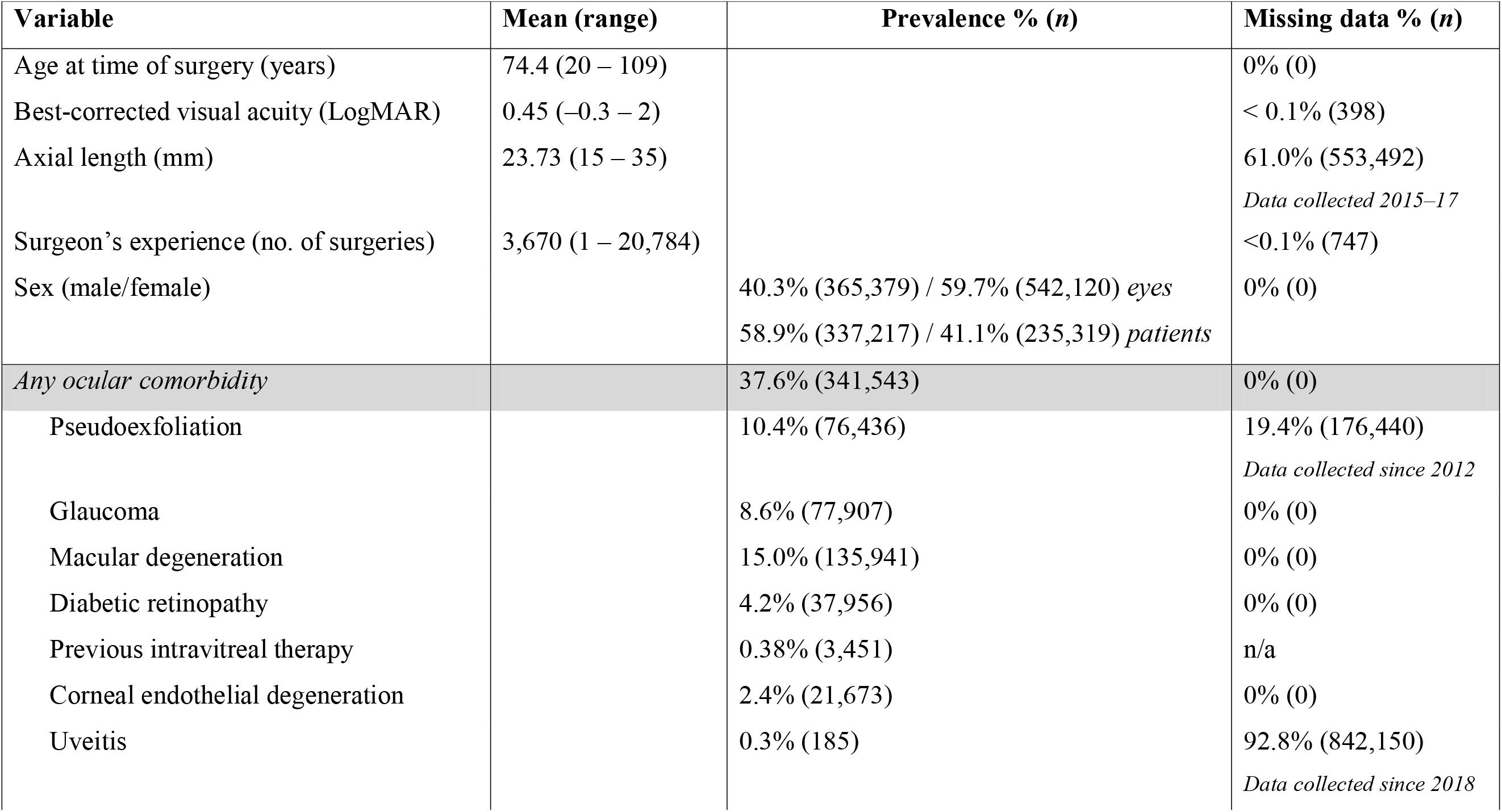
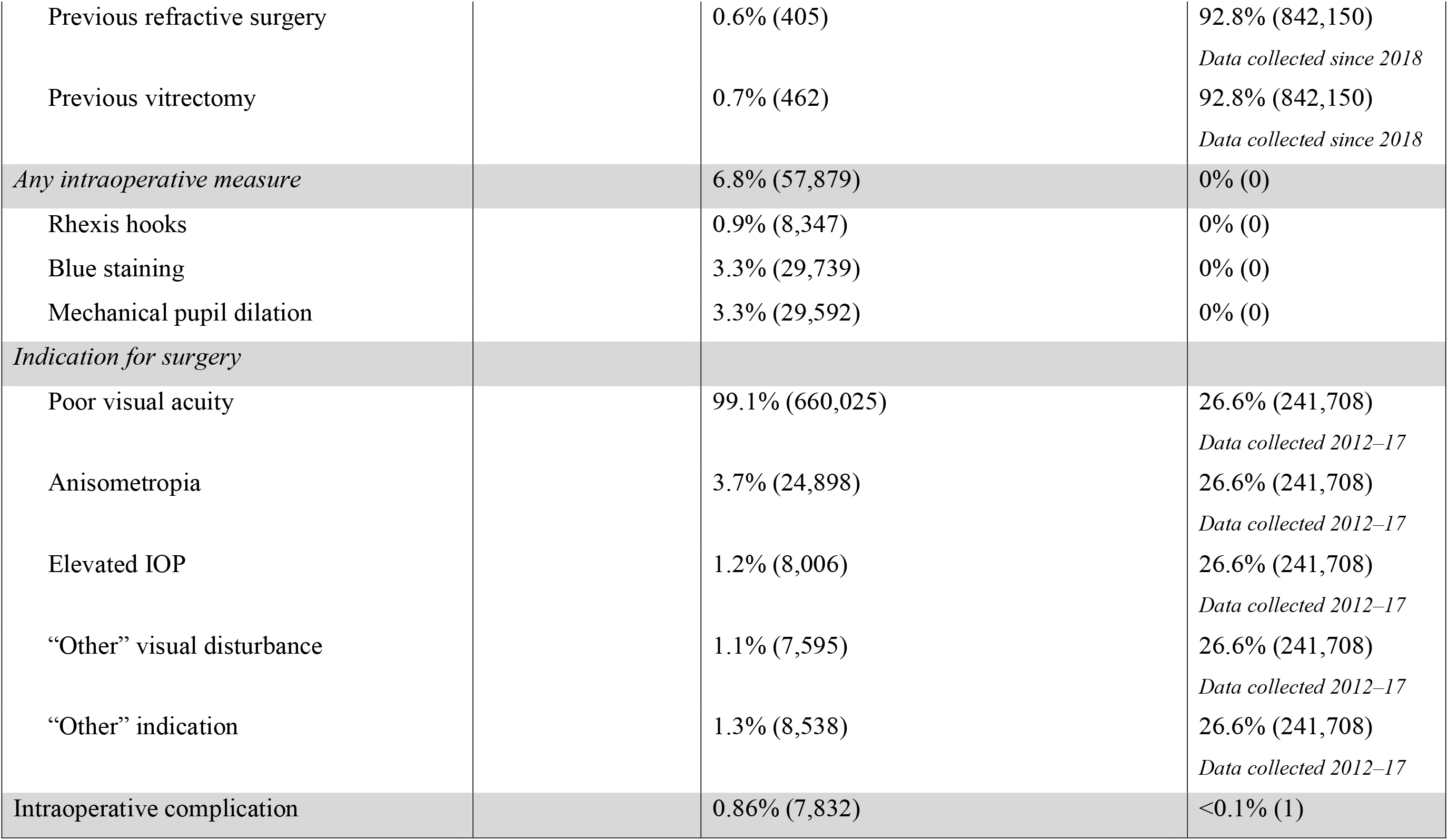
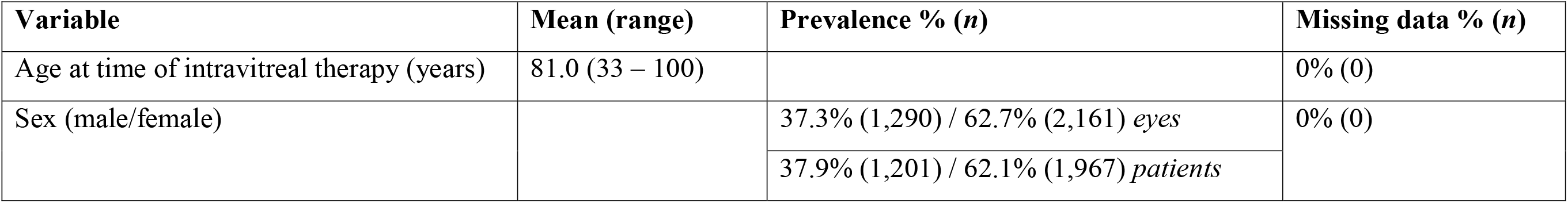
Characteristics of the patients identified in the Swedish National Cataract Register and the Swedish Macular Register. ‘Prevalence’ indicates the percentage (and number (*n*)) of eyes with valid data entries. ‘Missing data’ indicates the percentage (and number (*n*)) of eyes with missing data. a) Swedish National Cataract Register. Phacoemulsification cataract surgery data in 907,499 eyes in 572,536 patients >20 years age, according to the Swedish National Cataract Register for the period Jan. 1, 2010 – Jun. 30, 2018 (unless otherwise stated). b) Intravitreal therapy data in 3,451 eyes of 3,168 patients in the Swedish Macula Register found after cross referencing.

| Variable | Mean (range) | Prevalence % (*n*) | Missing data % (*n*) |
| --- | --- | --- | --- |
| Age at time of surgery (years) | 74.4 (20 – 109) |  | 0% (0) |
| Best-corrected visual acuity (LogMAR) | 0.45 (–0.3 – 2) |  | < 0.1% (398) |
| Axial length (mm) | 23.73 (15 – 35) |  | 61.0% (553,492)<br>*Data collected 2015–17* |
| Surgeon’s experience (no. of surgeries) | 3,670 (1 – 20,784) |  | <0.1% (747) |
| Sex (male/female) |  | 40.3% (365,379) / 59.7% (542,120) *eyes*<br>58.9% (337,217) / 41.1% (235,319) *patients* | 0% (0) |
| **Any ocular comorbidity** |  | **37.6% (341,543)** | **0% (0)** |
| Pseudoexfoliation |  | 10.4% (76,436) | 19.4% (176,440)<br>*Data collected since 2012* |
| Glaucoma |  | 8.6% (77,907) | 0% (0) |
| Macular degeneration |  | 15.0% (135,941) | 0% (0) |
| Diabetic retinopathy |  | 4.2% (37,956) | 0% (0) |
| Previous intravitreal therapy |  | 0.38% (3,451) | n/a |
| Corneal endothelial degeneration |  | 2.4% (21,673) | 0% (0) |
| Uveitis |  | 0.3% (185) | 92.8% (842,150)<br>*Data collected since 2018* |
| *Hård af Segerstad* |  |  |  |
| Previous refractive surgery |  | 0.6% (405) | 92.8% (842,150)<br>*Data collected since 2018* |
| Previous vitrectomy |  | 0.7% (462) | 92.8% (842,150)<br>*Data collected since 2018* |
| *Any intraoperative measure* |  | 6.8% (57,879) | 0% (0) |
| Rhexis hooks |  | 0.9% (8,347) | 0% (0) |
| Blue staining |  | 3.3% (29,739) | 0% (0) |
| Mechanical pupil dilation |  | 3.3% (29,592) | 0% (0) |
| *Indication for surgery* |  |  |  |
| Poor visual acuity |  | 99.1% (660,025) | 26.6% (241,708)<br>*Data collected 2012–17* |
| Anisometropia |  | 3.7% (24,898) | 26.6% (241,708)<br>*Data collected 2012–17* |
| Elevated IOP |  | 1.2% (8,006) | 26.6% (241,708)<br>*Data collected 2012–17* |
| “Other” visual disturbance |  | 1.1% (7,595) | 26.6% (241,708)<br>*Data collected 2012–17* |
| “Other” indication |  | 1.3% (8,538) | 26.6% (241,708)<br>*Data collected 2012–17* |
| Intraoperative complication |  | 0.86% (7,832) | <0.1% (1) |

| **Variable** | **Mean (range)** | **Prevalence % (*n*)** | **Missing data % (*n*)** |
| --- | --- | --- | --- |
| Age at time of intravitreal therapy (years) | 81.0 (33 – 100) |  | 0% (0) |
| Sex (male/female) |  | 37.3% (1,290) / 62.7% (2,161) *eyes* | 0% (0) |
|  |  | 37.9% (1,201) / 62.1% (1,967) *patients* |  |

**Figure 1.**
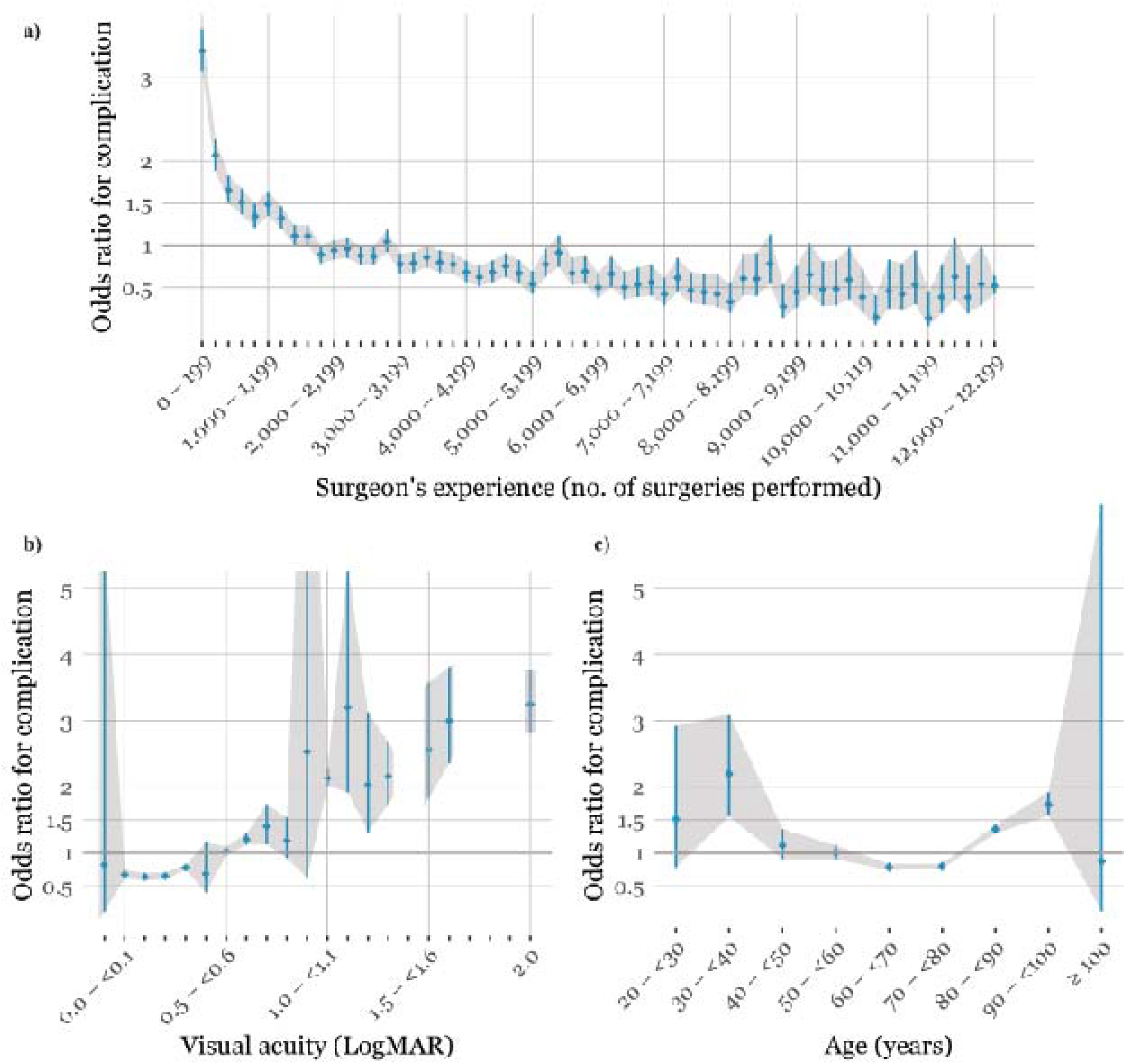
**a)** Odds ratio for complication as a function of the surgeon’s experience expressed as the number of surgeries performed, in intervals of 200. **b)** Odds ratio for complication as a function of BCVA in LogMAR, in intervals of 0.10. Empty areas in the graph indicate missing data due to an artefact after conversion from Snellen scores to LogMAR values. **c)** Odds ratio for complication as a function of patient age in years, in intervals of 10 years.

### Unadjusted risk factors

Patient characteristics associated with increased risk of IC were BCVA ≥1.0 LogMAR (corresponding to Snellen scores ≤ 0.1), age <40 years, age ≥90 years and male sex, together with elevated IOP and “other” indications for surgery. Ocular comorbidities associated with increased risk were previous vitrectomy, pIVT (dichotomized “yes” or “no”), pseudoexfoliation, glaucoma, and diabetic retinopathy. Previous intravitreal therapy was also calculated as the number of injections prior to cataract surgery; OR per injection: 1.03, 95% confidence interval (CI): 1.01–1.05 (*P* <0.01). Surgeon’s experience <600 surgeries was also associated with increased risk. Intraoperative difficulties were associated with the highest risk for IC: use of rhexis hooks, blue staining and mechanical pupil dilation, which indicate zonular instability, white cataract, and small pupil, respectively. To explore confounding effects between glaucoma and pseudoexfoliation, the univariate risk of IC was calculated after exclusion of pseudoexfoliation from glaucoma eyes. Glaucoma remained significant (OR: 1.49, 95% CI: 1.29–1.74, *P* <0.001). Anisometropia and low visual acuity were the only protective univariate factors. The ocular comorbidities macular degeneration, corneal endothelial degeneration, uveitis, previous refractive surgery, and the indication “other visual disturbance” were found to be nonsignificant. The results of the univariate analyses are summarized in Figure 2a.

**Figure 2.**
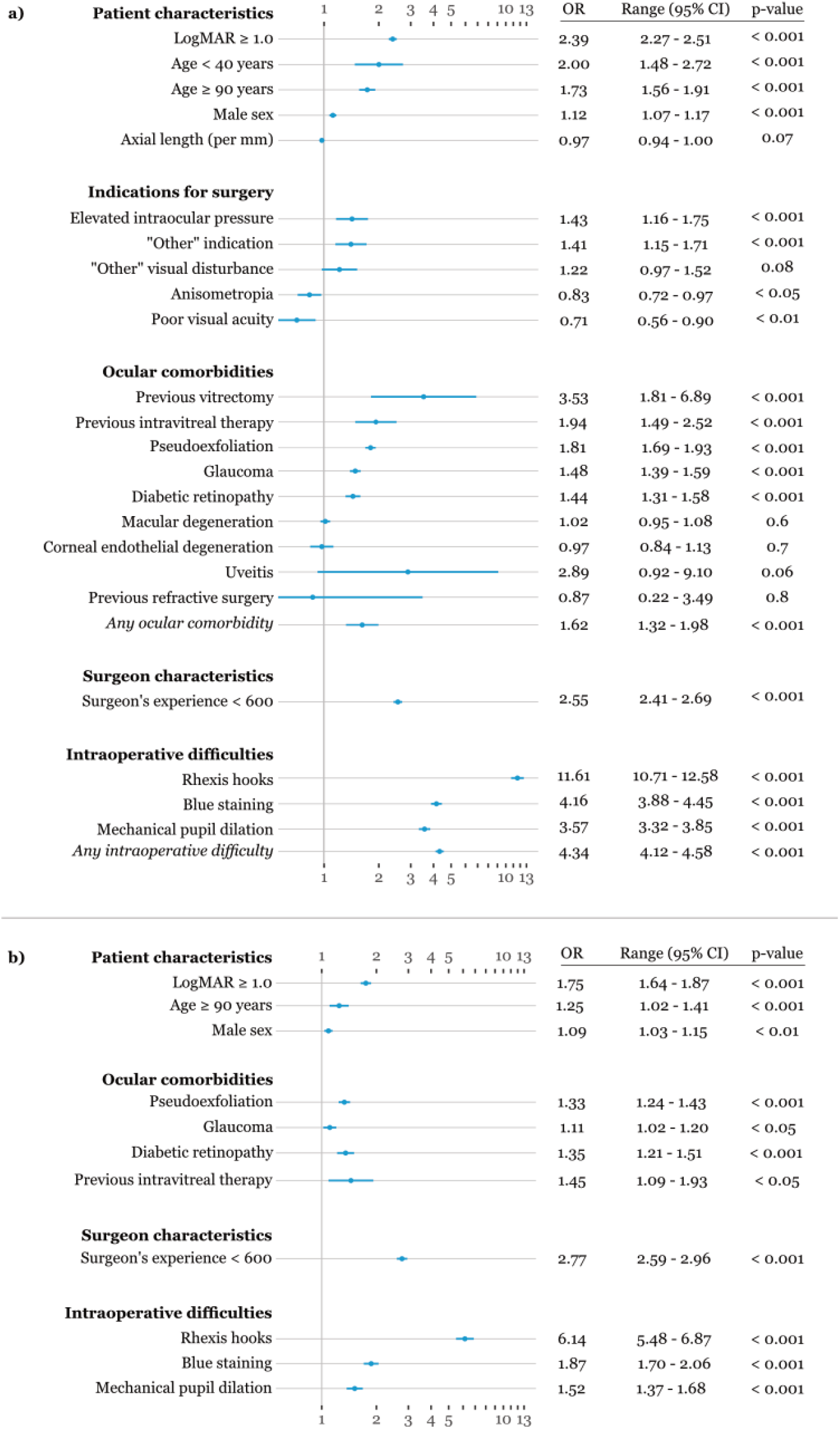
**a)** Unadjusted odds ratios of risk factors. Variables that cross the vertical line at a value of 1 are nonsignificant. Error bars indicate 95% confidence interval (95% CI). **b)** Adjusted odds ratios of risk factors in multivariate logistic regression model. Variables that cross the vertical line at 1 on the x-axis indicate nonsignificance. Error bars indicate 95% confidence interval. OR = odds ratio; 95% CI = 95% confidence interval.

### Adjusted risk factors

Variables found to be significant in the univariate analyses were included in a multivariate logistic regression model. Cases with missing data (177,179) were omitted, resulting in 730,320 eyes being included in the multivariate logistic regression model. All parameters remained significant except age group <40 years, and all indications for surgery. Previous vitrectomy could not be included due to the high number of missing data, and was excluded from the final model. Previous intravitreal therapy remained significant when included as the number of injections (OR per injection: 1.02, 95% CI:1.00–1.04, *P* <0.05). The results of the multivariate logistic regression analysis are summarized in Figure 2b.

In order to determine the probability of IC prior to surgery, the adjusted risk factors for a patient were combined by multiplication of the ORs of each variable, which resulted in an OR product. A similar approach has been used in previous studies^4^. This was converted in to the probability of complication by using the formula 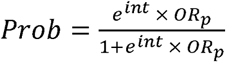; where the superscript *int* refers to the logistic regression intercept (–5.20), and *OR*_*p*_ denotes the odds ratio product. The equation is plotted in Figure 3. For example, a male patient aged over 89 years, with pseudoexfoliation, glaucoma, and a small pupil size who is believed to require mechanical pupil dilation has an odds ratio product of 3.06. This gives a predicted probability of IC of 1.7%. If this patient was operated on by a surgeon who had performed <600 surgeries, the odds ratio product would be 8.47 and the probability of an IC would be 4.5%. Using this model, the probability of ICs was calculated for each eye in the dataset, and was found to be less than 1.0% for 84% of the eyes in the dataset. As can be seen in Figure 4, the number of eyes declined exponentially with increasing risk. The mean and median risk of the dataset were 0.82 ± 0.99% and 0.56%, respectively. The mean probability of IC for eyes that were reported as having complication (*n*=6,001) was 1.85 ± 3.18%, compared with 0.81 ± 0.95% for eyes without complication (*n*=724,319).

**Figure 3.**
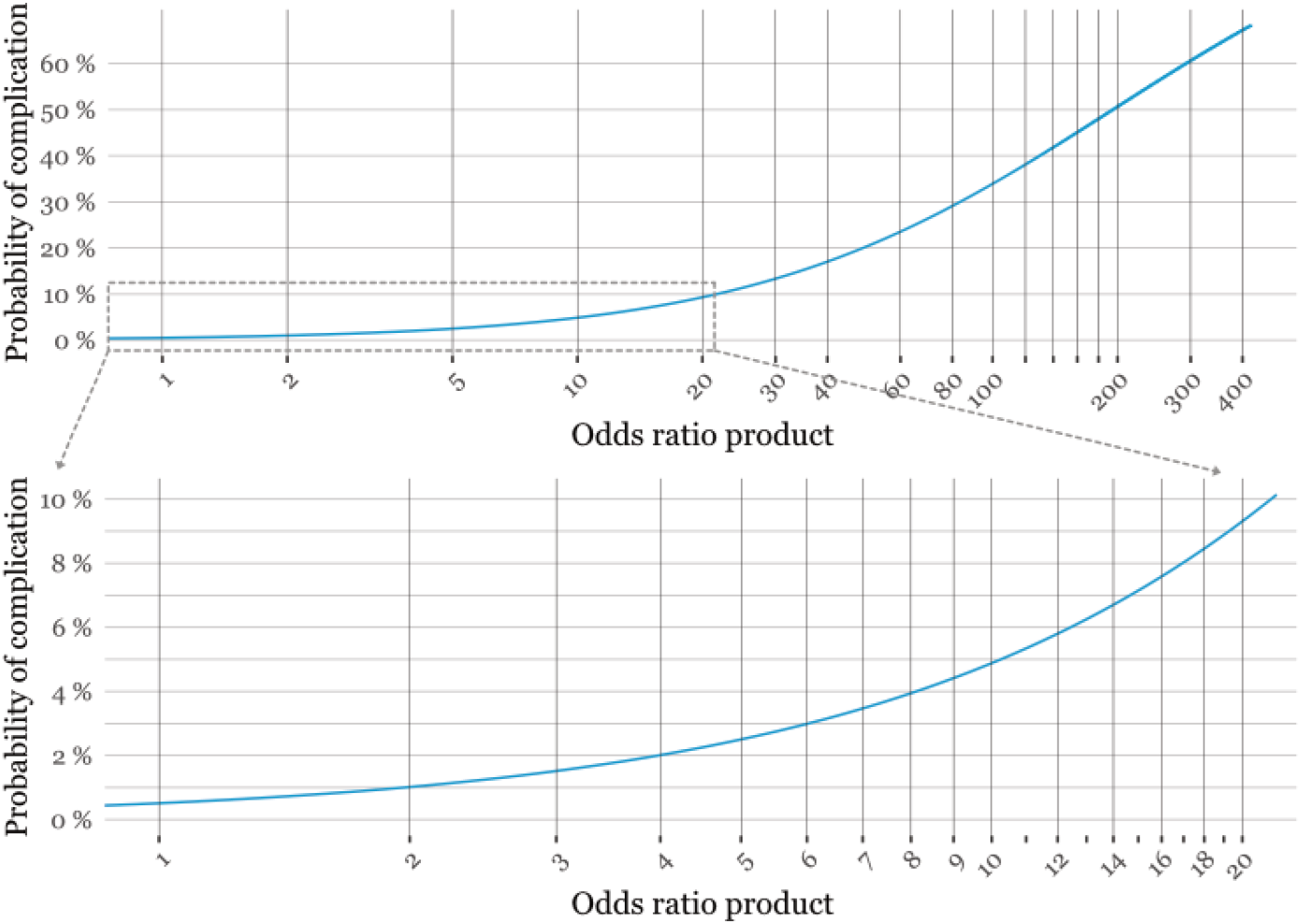
Probability (or risk) of complication as a function of the odds ratio product. The probability of complication can be read off the y-axis after calculating the product of the odds ratios.

**Figure 4.**
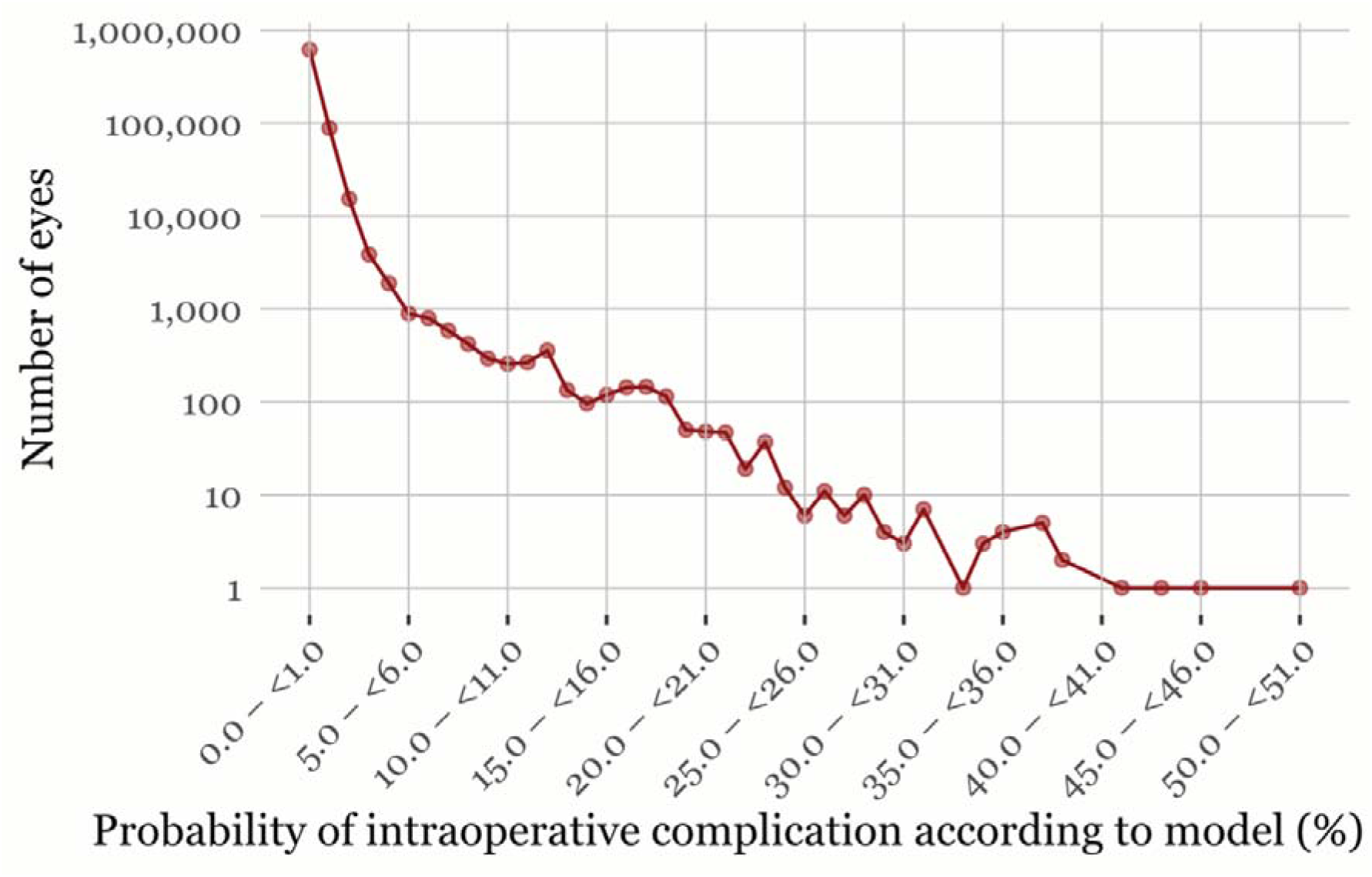
Frequency of the probability of intraoperative complication of the eyes in the dataset (note logarithmic y-scale). The risk of complications was low for most eyes.

## DISCUSSION

The main purpose of this retrospective database study was to develop a robust risk model for IC during cataract surgery and to investigate the impact of pIVT on the risk. In line with the findings of Lee et al.^7^ and Shalchi et al.^8^, it was found that pIVT was associated with an increased risk after adjusting for age, BCVA, sex, glaucoma, pseudoexfoliation, diabetic retinopathy, “other” ocular comorbidity, the surgeon’s experience, and intraoperative difficulties (rhexis hooks, blue staining and mechanical pupil dilation). Over 900,000 eyes reported to the SNCR from Jan. 1, 2010 to Jun. 30, 2018, were cross referenced with eyes reported to the SMR that had undergone intravitreal therapy prior to cataract surgery. Both registers have an exceptionally high degree of coverage^18, 19^. In our dataset, 0.38% (3,451 of 907,499 eyes) were treated for choroidal neovascularization before cataract surgery, which is in contrast to the prevalence reported in previous studies by Lee et al.^7^ and Shalchi et al.^8^, where the corresponding values were 2.9% and 1.6%, respectively. This could partly be explained by the fact that the SMR only includes eyes treated for choroidal neovascularization, while diabetes and other indications were included in the data used in the above-mentioned studies. Eyes previously subjected to intravitreal therapy were included in the present study irrespective of the number of injections. When investigating the correlation between the number of injections and the risk of ICs, both the univariate and multivariate analyses showed that a higher number of injections was associated with increased risk. Although it may be difficult to keep track of the number of injections, this information can be used in the model using *OR*^*n*^, where *OR* is the multivariate odds ratio per injection (OR: 1.02, 95% CI: 1.00–1.04, *P* <0.05), and *n* the number of previous intravitreal injections. Lee et al.^7^ also found that the number of injections was associated with increased risk. This is in contrast to the findings of Shalchi et al.^8^, who reported no association between the number of injections and the risk of complication.

Swedish personal ID number was introduced in the SNCR from 2010. Therefore, the data on eyes were extracted from Jan. 1, 2010 (to Jun. 30, 2018) except for surgeon’s experience. The surgeon’s experience was not a parameter collected in the SNCR, but was included in the analyses by counting the number of surgeries per surgeon from the year surgeon ID was introduced, Jan. 1, 2007. A limitation of this method was that all the surgeons started at “0” surgeries in 2007, regardless of their previous experience. Number of performed surgeries per surgeon was therefore calculated from Jan. 1, 2007 to Dec. 31, 2009 before being included in the eye dataset. By this approach, we eliminated the possibility that surgeons that gained experience during 2007–2009 would be assigned surgeon’s experience “0”. Due to the high coverage rate of the SNCR, surgeons having “0” surgeries in the dataset were most likely beginners. The risk of IC was seen to decrease significantly as a result of gained experience, and reached a steady value below the odds ratio of 1.0 after about 1,800 surgeries. This was in line with Böhringer et al.^20^ who reported a decreasing rate of posterior capsule rupture from 4% in novice surgeons (with less experience than 300 procedures), to 0.5%–1% in surgeons with more than 1,500 procedures experience.

The patient’s age, and BCVA at which the risk of IC was significantly affected (odds ratio >1.5) were in line with those reported in some previous studies^4, 21, 22^. Narendran et al.^4^ reported the odds ratio for posterior capsule rupture, vitreous loss or both to be 2.18 in patients aged above 90 years. Zetterberg et al.^21^ reported the odds ratio for capsule complication to be 1.82 in eyes with BCVA <0.1 Snellen (>1.0 LogMAR), while Theodoropoulou^22^ reported an odds ratio of 2.19 in eyes with BCVA >1.2 LogMAR. In the present study, no specific cut-off value was found for axial length (data not shown). This is believed to be mainly due to the chosen interval of 1.0 mm of axial length when calculating the odds ratios for IC. It is possible that comparing odds ratios for larger intervals of axial length than 1.0 mm might have given different results.

Intraoperative complication occurred in 0.83% (6,001 of 724,319 eyes) of all reported cataract surgeries. This is over a factor of ten higher than the value of 0.071% (1,199 of 1,687,635 eyes) reported by Lundström et al.^3^ using data from the European Registry of Quality Outcomes for Cataract and Refractive Surgery (EUREQUO). The reason for the considerable difference in reported complications between the Swedish and European data is not clear, however, there is a difference in the way in which the data are reported. The SNCR includes data from almost all cataract surgeries in Sweden, while the EUREQUO is based on voluntary reporting, which may lead to incomplete coverage and lower rates of reported complications.

The same approach as used by Zetterberg et al.^21^ was used to control for confounding effects between glaucoma and pseudoexfoliation. Glaucoma was found to be significant in the present study, which is in contrast to their findings. This is believed to be due to the higher numbers of eyes in the present study. As odds ratios are difficult to interpret and seldom used in the clinical setting, they were converted to probability, which is easier to understand and communicate. The single eye with the highest probability of ICs in the present study was that of a female aged less than 90 years, with a BCVA greater than 1.0 LogMAR, glaucoma, and diabetic retinopathy, operated on by an unexperienced surgeon (<600 surgeries) using rhexis hooks, blue staining, and mechanical pupil dilation. The probability of IC in this eye was 51%, but no IC was actually reported. The eye with the theoretically highest risk would yield a complication probability of 71%.

The results of this study can be used in planning cataract surgery, and to improve communication with the patient prior to surgery. Patients with a probability of complications above a certain level could be allocated to more experienced cataract surgeons, and those with the highest risk to cataract surgeons with vitreoretinal competency, so that capsule complication and lens implantation could be addressed during the same session. The thresholds used to categorize patients according to risk will depend on the demographics and resources of individual healthcare providers. The strengths of this study lie in the high number of eyes and the high coverage rate of the unique nationwide registers, while previous studies have relied on local databases. In addition, the combination of risk factors in this study have not been not presented before, in particular the use of previous intravitreal therapy data from the Swedish Macula Register together with the commonly known factors such as pseudoexfoliation. Moreover, the span of more than eight years data collection contributes to the robustness of the model, while the translation from odds ratios to probability of intraoperative complication enables use of the model in a clinical setting prior to surgery. One of the limitations is the selection of variables reported to the registers. For instance, IC is defined only as “communication between the anterior and posterior segment”, meaning posterior capsule rupture, or dropped nucleus. However, other types of complication can occur intraoperatively, such as wound complications, corneal edema, iris prolapse, iris bleeding, anterior capsule complications, and expulsive bleeding, which are not included in the definition. Also, some of the parameters cannot always be foreseen preoperatively, such as the use of rhexis hooks, blue staining, or mechanical pupil dilation, although they are considered to be proxy markers for zonular instability, white cataract, and small pupil. The use of these measures is highly dependent on the surgeon; some rarely use rhexis hooks, while others use blue staining routinely in the early phases of training. However, the number of parameters recorded in registers should be restricted to ensure easy reporting and high coverage. Furthermore, errors in entering the data, misunderstandings, and miscommunication are inevitable in the reporting process, and the data used in this study had not been verified against the patients’ medical records. This issue could be addressed by direct reporting from the medical record to the different registers in the future.

In conclusion, it was found that pIVT increased the risk of IC during cataract surgery, based on data from 907,499 eyes reported to the SNCR from January 1, 2010–June 30, 2018. A risk model was also presented that can be used to quantify the probability of IC. The application of this model in the clinical setting prior to surgery allows patients to be allocated to surgeons with the appropriate level of experience and competence, as well as improving communication with the patient regarding the risks involved.

## Data Availability

The data is available for use in clinical research after ethical approval.

## ACKNOWLEDGMENTS

The author thanks Mr. Axel Ström, M.Sc., Forum Söder, for extensive statistical assistance. This study was partly supported by the Foundation for Visually Impaired in former Malmöhus Län, The Eye Foundation (Ögonfonden) and The Cronqvist Foundation. The funding organizations had no role in the design or conduct of this research.

## REFERENCES

1. Hashemi H, Pakzad R, Yekta A, et al. Global and regional prevalence of age-related cataract: a comprehensive systematic review and meta-analysis. Eye (Lond). Aug 2020;34(8):1357–1370. doi:10.1038/s41433-020-0806-3

2. Chen M, Lamattina KC, Patrianakos T, Dwarakanathan S. Complication rate of posterior capsule rupture with vitreous loss during phacoemulsification at a Hawaiian cataract surgical center: a clinical audit. Clin Ophthalmol. 2014;8:375–8. doi:10.2147/OPTH.S57736

3. Lundström M, Dickman M, Henry Y, et al. Risk factors for dropped nucleus in cataract surgery as reflected by the European Registry of Quality Outcomes for Cataract and Refractive Surgery. J Cataract Refract Surg. 02 2020;46(2):287–292. doi:10.1097/j.jcrs.0000000000000019

4. Narendran N, Jaycock P, Johnston RL, et al. The Cataract National Dataset electronic multicentre audit of 55,567 operations: risk stratification for posterior capsule rupture and vitreous loss. Eye (Lond). Jan 2009;23(1):31–7. doi:10.1038/sj.eye.6703049

5. Ionides A, Minassian D, Tuft S. Visual outcome following posterior capsule rupture during cataract surgery. Br J Ophthalmol. Feb 2001;85(2):222–4. doi:10.1136/bjo.85.2.222

6. Lundström M, Behndig A, Kugelberg M, Montan P, Stenevi U, Thorburn W. Decreasing rate of capsule complications in cataract surgery: eight-year study of incidence, risk factors, and data validity by the Swedish National Cataract Register. J Cataract Refract Surg. Oct 2011;37(10):1762–7. doi:10.1016/j.jcrs.2011.05.022

7. Lee AY, Day AC, Egan C, et al. Previous Intravitreal Therapy Is Associated with Increased Risk of Posterior Capsule Rupture during Cataract Surgery. Ophthalmology. Jun 2016;123(6):1252–6. doi:10.1016/j.ophtha.2016.02.014

8. Shalchi Z, Okada M, Whiting C, Hamilton R. Risk of posterior capsule rupture during cataract surgery in eyes with previous intravitreal injections. Am J Ophthalmol. Feb 14 2017;doi:10.1016/j.ajo.2017.02.006

9. Khanna S, Komati R, Eichenbaum DA, Hariprasad I, Ciulla TA, Hariprasad SM. Current and upcoming anti-VEGF therapies and dosing strategies for the treatment of neovascular AMD: a comparative review. BMJ Open Ophthalmol. 2019;4(1):e000398. doi:10.1136/bmjophth-2019-000398

10. Cedrone C, Culasso F, Cesareo M, et al. Prevalence and incidence of age-related cataract in a population sample from Priverno, Italy. Ophthalmic Epidemiol. Jun 1999;6(2):95–103. doi:10.1076/opep.6.2.95.1562

11. Congdon N, Vingerling JR, Klein BE, et al. Prevalence of cataract and pseudophakia/aphakia among adults in the United States. Arch Ophthalmol. Apr 2004;122(4):487–94. doi:10.1001/archopht.122.4.487

12. Rein DB, Wittenborn JS, Zhang X, et al. Forecasting age-related macular degeneration through the year 2050: the potential impact of new treatments. Arch Ophthalmol. Apr 2009;127(4):533–40. doi:10.1001/archophthalmol.2009.58

13. Behndig A, Montan P, Stenevi U, Kugelberg M, Lundstrom M. One million cataract surgeries: Swedish National Cataract Register 1992-2009. J Cataract Refract Surg. Aug 2011;37(8):1539–45. doi:10.1016/j.jcrs.2011.05.021

14. Westborg I, Albrecht S, Granstam E, et al. Treatment of age-related macular degeneration after cataract surgery: a study from the Swedish National Cataract and Macula Registers. Acta Ophthalmol. Jun 2020;doi:10.1111/aos.14519

15. Westborg I, Granstam E, Rosso A, Albrecht S, Karlsson N, Lövestam-Adrian M. Treatment for neovascular age-related macular degeneration in Sweden: outcomes at seven years in the Swedish Macula Register. Acta Ophthalmol. Dec 2017;95(8):787–795. doi:10.1111/aos.13539

16. Holladay JT. Proper method for calculating average visual acuity. J Refract Surg. 1997 Jul-Aug 1997;13(4):388–91.

17. R Code Team R: A Language and Environment for Statistical Computing,. 2018Located at: R Foundation for Statistical Computing, Vienna, Austria.

18. Swedish National Cataract Register Annual Reports 2010 -2018. Accessed November, 2020. http://rcsyd.se/kataraktreg/publikationer/arsrapporter

19. Swedish Macula Register Annual Reports 2010 - 2018. Accessed November, 2020. http://makulareg.se/publikationer/arsrapporter

20. Böhringer D, Vach W, Hagenlocher K, Eberwein P, Maier P, Reinhard T. Are entry criteria for cataract surgery justified? PLoS One. 2014;9(11):e112819. doi:10.1371/journal.pone.0112819

21. Zetterberg M, Kugelberg M, Nilsson I, Lundström M, Behndig A, Montan P. A Composite Risk Score for Capsule Complications Based on Data from the Swedish National Cataract Register: Relation to Surgery Volumes. Ophthalmology. Jul 2020;doi:10.1016/j.ophtha.2020.07.033

22. Theodoropoulou S, Grzeda MT, Donachie PHJ, Johnston RL, Sparrow JM, Tole DM. The Royal College of Ophthalmologists’ National Ophthalmology Database Study of cataract surgery. Report 5: Clinical outcome and risk factors for posterior capsule rupture and visual acuity loss following cataract surgery in patients aged 90 years and older. Eye (Lond). 07 2019;33(7):1161–1170. doi:10.1038/s41433-019-0389-z

